# Assessing Differential Impacts of COVID-19 on Black Communities

**DOI:** 10.1101/2020.05.04.20090274

**Authors:** Gregorio A. Millett, Austin T. Jones, David Benkeser, Stefan Baral, Laina Mercer, Chris Beyrer, Brian Honermann, Elise Lankiewicz, Leandro Mena, Jeffrey S. Crowley, Jennifer Sherwood, Patrick Sullivan

## Abstract

**Purpose:** Given incomplete data reporting by race, we used data on COVID-19 cases and deaths in US counties to describe racial disparities in COVID-19 disease and death and associated determinants.

**Methods:** Using publicly available data (accessed April 13, 2020), predictors of COVID-19 cases and deaths were compared between disproportionately (≥13%) black and all other (<13% black) counties. Rate ratios were calculated and population attributable fractions (PAF) were estimated using COVID-19 cases and deaths via zero-inflated negative binomial regression model. National maps with county-level data and an interactive scatterplot of COVID-19 cases were generated.

**Results:** Nearly ninety-seven percent of disproportionately black counties (656/677) reported a case and 49% (330/677) reported a death versus 81% (1987/2,465) and 28% (684/ 2465), respectively, for all other counties. Counties with higher proportions of black people have higher prevalence of comorbidities and greater air pollution. Counties with higher proportions of black residents had more COVID-19 diagnoses (RR 1.24, 95% CI 1.17-1.33) and deaths (RR 1.18, 95% CI 1.00-1.40), after adjusting for county-level characteristics such as age, poverty, comorbidities, and epidemic duration. COVID-19 deaths were higher in disproportionally black rural and small metro counties. The PAF of COVID-19 diagnosis due to lack of health insurance was 3.3% for counties with <13% black residents and 4.2% for counties with ≥13% black residents.

**Conclusions:** Nearly twenty-two percent of US counties are disproportionately black and they accounted for 52% of COVID-19 diagnoses and 58% of COVID-19 deaths nationally. County-level comparisons can both inform COVID-19 responses and identify epidemic hot spots. Social conditions, structural racism, and other factors elevate risk for COVID-19 diagnoses and deaths in black communities.

## Introduction

As of April 30, 2020, more than one million cases of severe acute respiratory syndrome coronavirus 2 (SARS-CoV-2) infection have been diagnosed in the United States, and deaths exceed 63,000.^1^ Emerging evidence suggests that black Americans are at increased risk for COVID-19 morbidity and mortality. Although it may be counter-intuitive that a newly identified virus that can infect anyone would rapidly manifest pronounced racial disparities, a consistent pattern has been reported across multiple states, showing that black Americans comprise a disproportionately greater number of reported COVID-19 cases and deaths compared to other Americans^2-4^ For instance, in New York City, the current epicenter of the US epidemic, COVID-19 deaths disproportionately affect black Americans (22% of population and 28% of deaths) and in the rest of the state (9% of the population and 18% of deaths).^5^ Such disparities are also evident within individual counties, such as in Milwaukee County, Wisconsin where black residents comprise 26% of the population yet account for 73% of COVID-19 deaths;^6^ and in Dougherty County, GA (69% black) where 81% of 38 deaths were black.^7^

Although the CDC reports cumulative COVID-19 data reported by state health departments, 78% of those data were missing race/ethnicity disaggregations as of April 15, 2020.^8^ Incomplete reporting of race remains common. One state’s department of health reported 20.5% of COVID-19 cases among blacks, 15.3% among whites, 1.5% other, yet nearly two-thirds (62.7%) were racial/ethnic category unknown.^9^ Another disaggregated their data, but reported that three quarters (74%) of the cases were of unknown race or ethnicity.^10^

A more thorough understanding of the impact of COVID-19 by race/ethnicity will remain unavailable until more states report disaggregated data and until additional work is conducted to strengthen the completeness of race/ethnicity data. In the meantime, it is possible to use ecological analyses with data at the county level to assess determinants of risk among black Americans to inform immediate policy actions. However, this approach risks confounding of the relationship between proportion of black Americans and disease and death rates in a county and other social determinants of health potentially associated with risk for COVID-19 infection and death, such as sociodemographic, comorbidities, and socioeconomic determinants.

We analyzed county-level data comparing counties with higher and lower proportions of black people to document whether COVID-19 diagnoses and deaths were higher in counties with higher proportions of black Americans. In addition, we used multivariable modeling to assess whether observed disproportionate impacts of COVID-19 disease and death in disproportionately black counties were explained by confounding with comorbidities, social, and environmental factors? Finally, we calculated the population attributable fraction of COVID-19 diagnoses and deaths in disproportionately black counties that was associated with demographics (unemployment, uninsurance), comorbidities, social or environmental factors.

## Materials and Methods

U.S. counties were stratified by the population of black Americans nationally, as previously described.^11^ We assessed differences between in the characteristics of counties with a greater share of black residents than the US average (>13% black population; hereafter disproportionately black counties) versus all other counties (<13% black population). We subsequently examined associations between the proportion of black residents and COVID-19 cases and deaths. All data used in these analyses are from publicly available datasets.

### Demographic data

County-level data from the US Census Bureau American Community Survey 5-Year^12^ were collected for select demographics (i.e. county population, percent black American, percent of the population over the age of 65, percent of the under 65 population without health insurance, occupants per room). Annual average county unemployment rates were obtained from the Bureau of Labor Statistics.^13^

### COVID-19 data and co-morbidities

COVID-19 cases and deaths at the county-level were downloaded from USAFacts through April 13^th^.^14^ Rates of diagnosed diabetes among adults aged 20+ were downloaded from CDC Diabetes Atlas (2016).^15^ Heart disease per 100,000 were accessed from CDC’s Interactive Atlas of Heart Disease and Stroke (2014-16)^16^, and the combined rate of cerebrovascular and hypertension deaths per 100,000 were sourced from CDC WONDER (2018).^17^ Estimates of people living with diagnosed HIV per 100,000 among adults and adolescents 13 and older were derived from CDC ATLAS (2017).^18^ For counties with missing HIV data in Kentucky and Alaska, archived HIV data from AIDSVu.org^19^ from 2015 were used given that state data sharing agreements for these two states restrict CDC from releasing comparable data.

### Social/Environmental data

Following Wu, we use county-level estimates of fine particulate matter (PM2.5) to assess air quality.^20^ Social distancing grades were drawn from Unacast’s county measures on April 13 and were coded as A+/- = 1; B+/- = 2; C+/-=3; D+/-=4; F+/-=5.^21^ Thus, higher scores are associated with poorer social distancing. The CDC’s National Center for Health Statistics Urban-Rural Classification Scheme was used to assess urbanicity (index from 1-6, with 1 being the most urban).^22^

### Statistical Analyses

We compared characteristics of counties based on the proportion of the population who are black (< or ≥13%) using medians and interquartile ranges. We plotted the county proportion of black residents by the county rate of COVID-19 diagnoses, adjusted for days since first infection in the county. To assess whether observed associations between proportion of black residents and COVID-19 cases and deaths were confounded by other factors, we used Bayesian hierarchical models. A zero-inflated negative binomial model with a logarithmic link function was fit separately to COVID-19 cases and deaths using integrated nested Laplace approximations.^23^ The model included county population as an offset term, adjusted for all county-level characteristics (see Table 1 for characteristics), and included a spatially structured state-level random effect.^24^ We also included a variable representing days since the first case of COVID-19 was reported in each county in order to control for potential confounding by temporality of the outbreak. Diffuse priors were included for all components of the model. Exponentiated regression coefficients are presented along with 95% confidence intervals, representing rate ratios of COVID-19 cases and deaths. The primary question was whether the rate ratios COVID-19 cases and deaths for disproportionately black counties remained significant after controlling for sociodemographics, comorbidities, and socioeconomic determinants. In reporting results for the covariates, county-level characteristics are scaled to represent a comparison of two counties similar in all other respects, but for one county having the variable of interest equal to the observed third quartile of that characteristic, the other equal to the observed first quartile. Thus, rate ratios greater than one mean that higher levels of a given characteristic are associated with higher rates of COVID-19 cases or deaths. For “modifiable” risk factors (e.g., insurance, population density), we computed population attributable fractions by computing the multiplicative reduction in predicted number of cases/deaths under the observed data versus when all counties in the upper three quartiles of a risk factor were reduced to the first quartile. See online appendix for details.

**Table 1.**
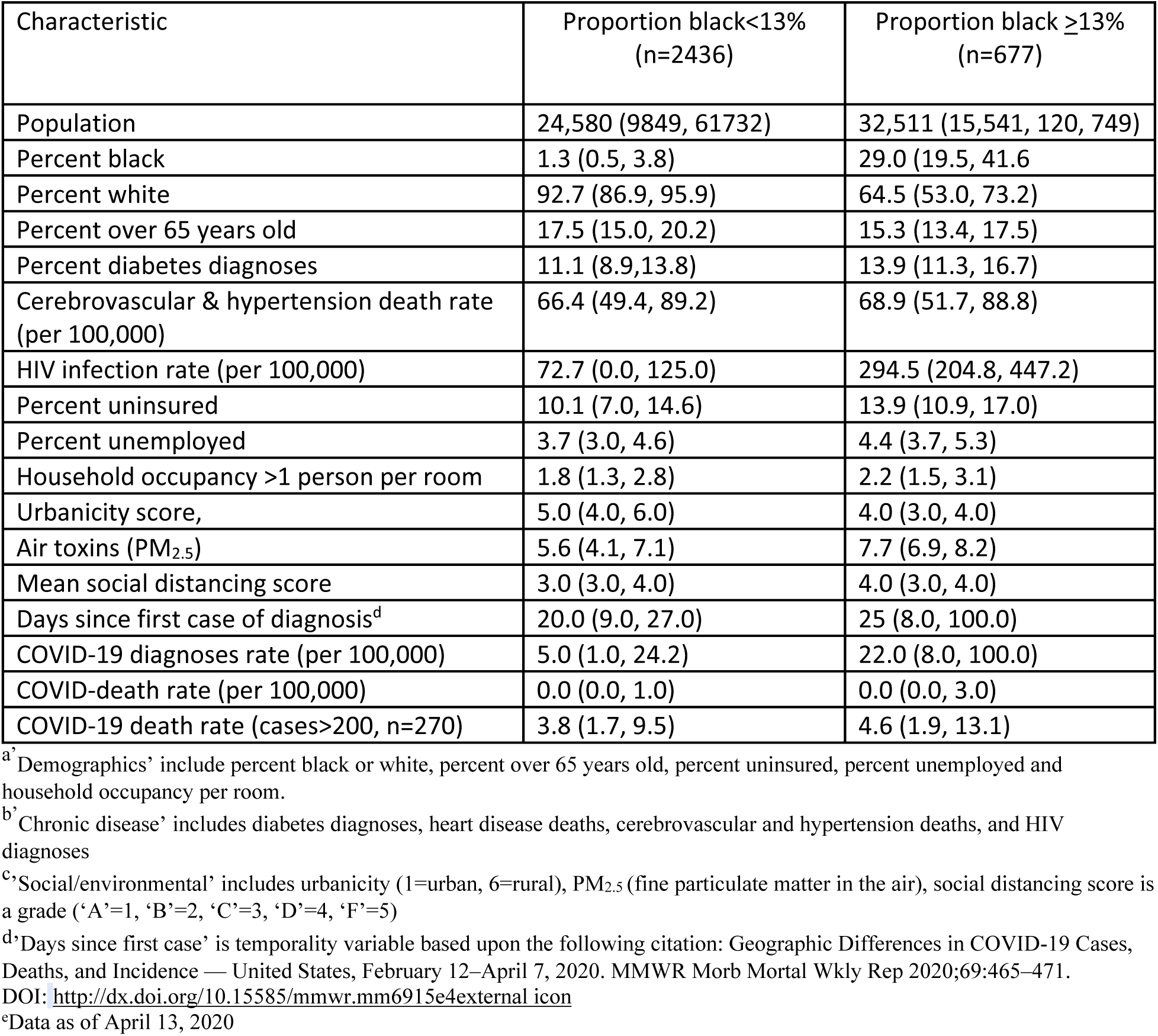
Unadjusted Associations of Demographics^a^, Chronic Disease^b^, Social/Environmental^c^ Factors, and COVID-19 Diagnoses and Deaths by Proportion of Black Residents

| Characteristic | Proportion black<13%<br>(n=2436) | Proportion black ≥13%<br>(n=677) |
| --- | --- | --- |
| Population | 24,580 (9849, 61732) | 32,511 (15,541, 120, 749) |
| Percent black | 1.3 (0.5, 3.8) | 29.0 (19.5, 41.6) |
| Percent white | 92.7 (86.9, 95.9) | 64.5 (53.0, 73.2) |
| Percent over 65 years old | 17.5 (15.0, 20.2) | 15.3 (13.4, 17.5) |
| Percent diabetes diagnoses | 11.1 (8.9,13.8) | 13.9 (11.3, 16.7) |
| Cerebrovascular & hypertension death rate<br>(per 100,000) | 66.4 (49.4, 89.2) | 68.9 (51.7, 88.8) |
| HIV infection rate (per 100,000) | 72.7 (0.0, 125.0) | 294.5 (204.8, 447.2) |
| Percent uninsured | 10.1 (7.0, 14.6) | 13.9 (10.9, 17.0) |
| Percent unemployed | 3.7 (3.0, 4.6) | 4.4 (3.7, 5.3) |
| Household occupancy >1 person per room | 1.8 (1.3, 2.8) | 2.2 (1.5, 3.1) |
| Urbanicity score, | 5.0 (4.0, 6.0) | 4.0 (3.0, 4.0) |
| Air toxins (PM<sub>2.5</sub>) | 5.6 (4.1, 7.1) | 7.7 (6.9, 8.2) |
| Mean social distancing score | 3.0 (3.0, 4.0) | 4.0 (3.0, 4.0) |
| Days since first case of diagnosis<sup>d</sup> | 20.0 (9.0, 27.0) | 25 (8.0, 100.0) |
| COVID-19 diagnoses rate (per 100,000) | 5.0 (1.0, 24.2) | 22.0 (8.0, 100.0) |
| COVID-death rate (per 100,000) | 0.0 (0.0, 1.0) | 0.0 (0.0, 3.0) |
| COVID-19 death rate (cases>200, n=270) | 3.8 (1.7, 9.5) | 4.6 (1.9, 13.1) |

<sup>a</sup>Demographics' include percent black or white, percent over 65 years old, percent uninsured, percent unemployed and household occupancy per room.
<sup>b</sup>Chronic disease' includes diabetes diagnoses, heart disease deaths, cerebrovascular and hypertension deaths, and HIV diagnoses
<sup>c</sup>'Social/environmental' includes urbanicity (1=urban, 6=rural), PM<sub>2.5</sub> (fine particulate matter in the air), social distancing score is a grade ('A'=1, 'B'=2, 'C'=3, 'D'=4, 'F'=5)
<sup>d</sup>'Days since first case' is temporality variable based upon the following citation: Geographic Differences in COVID-19 Cases, Deaths, and Incidence — United States, February 12–April 7, 2020. MMWR Morb Mortal Wkly Rep 2020;69:465–471.
DOI: <http://dx.doi.org/10.15585/mmwr.mm6915e4external icon>
<sup>e</sup>Data as of April 13, 2020

To evaluate the possibility of structural confounding^25,26^ (i.e., that higher rates of COVID-19 disease and death in disproportionately black counties might be driven by urban counties, which were impacted earlier by the epidemic and were more likely to be disproportionately black), we evaluated stratum-specific risk ratios associated with disproportionately black counties by level of urbanicity.

All analyses were performed using R 3.6.1. Bayesian hierarchical models were fit using the inla package (www.r-inla.org). All data and code needed to replicate the analysis are available online (https://github.com/benkeser/covid-and-race). There was no COVID-19 county-level data for Puerto Rico, and no PM_2.5_ data for Alaska. Both localities were excluded from the analysis.

## Results

Of 3,142 counties included in the analysis, 677 were disproportionately black. Ninety-seven percent (656/677) and 49% (330/677) of disproportionately black counties reported at least one COVID case and death, respectively. Eighty percent (1987/2,465) and 28% (684/2,465) of all other counties reported at least one COVID case and death, respectively. Ninety-one percent (616/677) of disproportionately black counties are located in the southern US (Figure 1). The proportion of black residents across disproportionately black counties ranged from 13.0% to 87.4%. As of April 13, 2020, there were 283,750 diagnoses in disproportionately black counties and 12,748 deaths. By comparison, all other counties had 263,640 diagnoses and 8,886 deaths. Collectively, 52% of COVID-19 cases and 58% of COVID-19 deaths occurred in disproportionally black counties.

**Figure 1.**
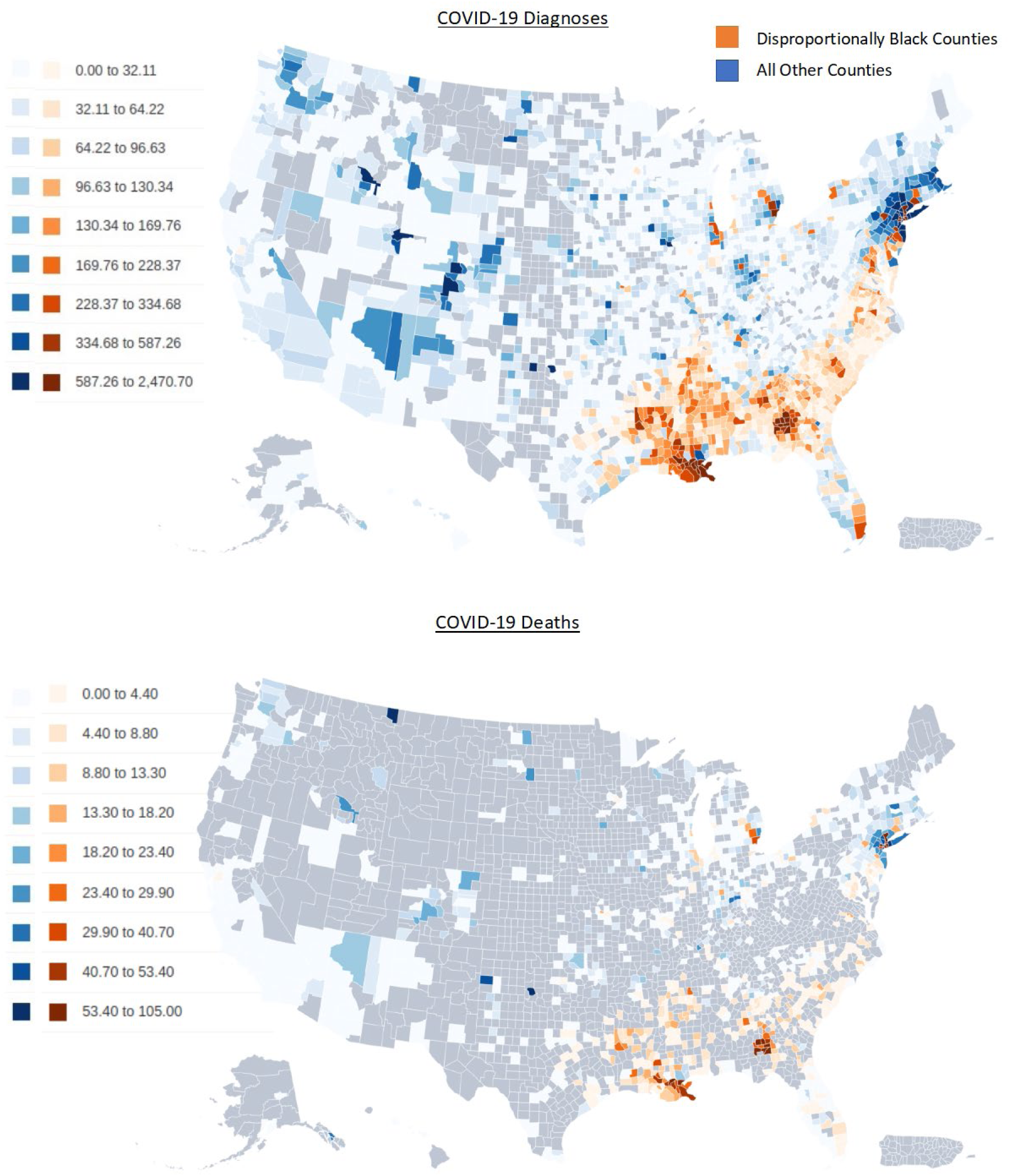
Rates of COVID-19 Diagnoses and Deaths in Disproportionately Black (≥13% of population) Versus All Other Counties (<13% of population) as of April 13, 2020

Figure 2 plots COVID-19 cases per 100,000 population (adjusted for days since detection) by proportion of black residents. (An interactive plot with ‘hover over’ data display functionality is available at https://ehe.amfar.org/bubble_plot_cases.html). For counties with ≥5% of black residents, higher proportions of black residents were associated with higher rates of COVID-19 diagnoses.

**Figure 2.**
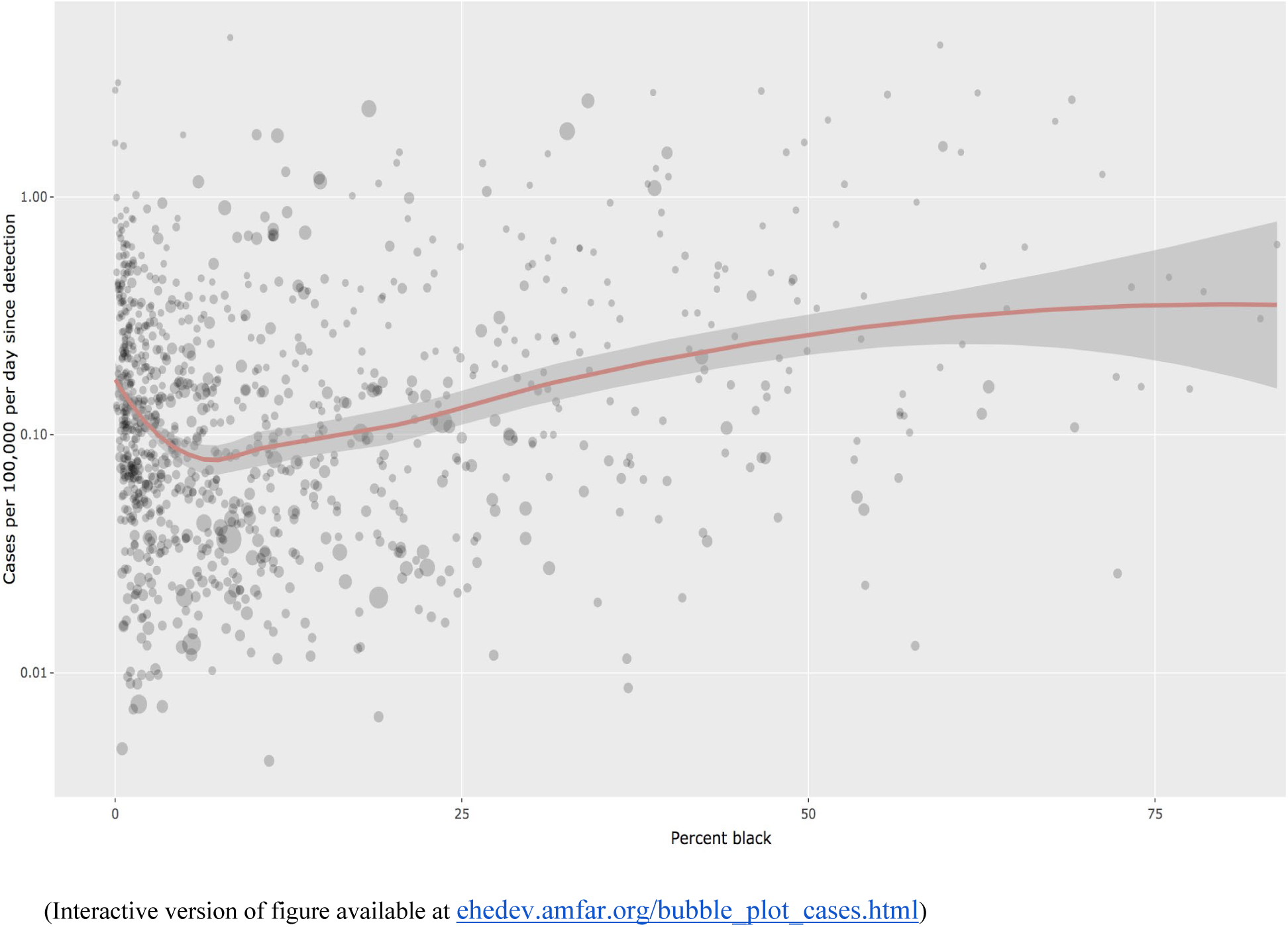
COVID-19 Cases Per 100,000 (adjusted per day since detection) by Increasing Proportion of Black Residents Across US Counties as of April 13, 2020

**Figure 3.**
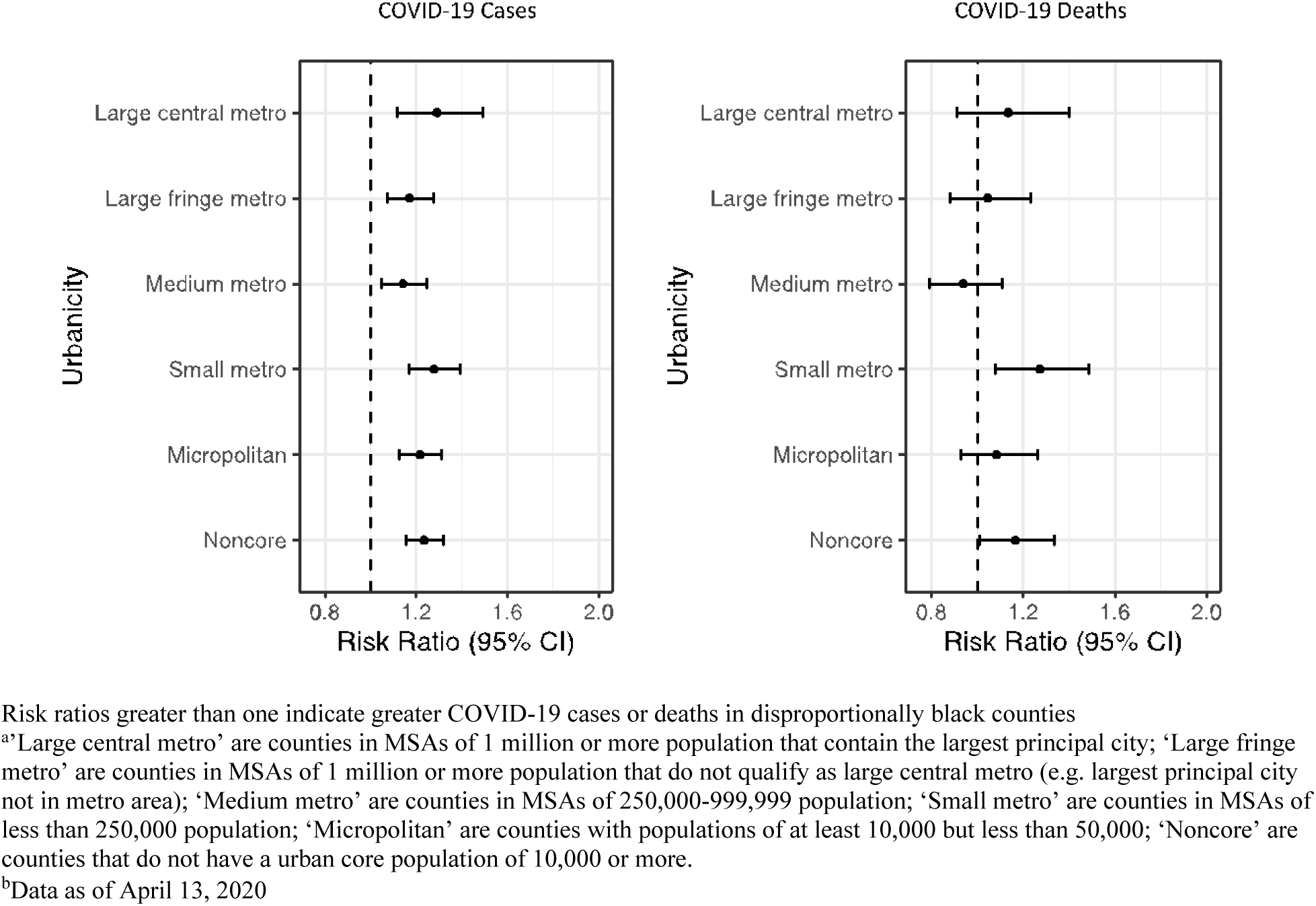
Forest Plots of COVID-19 cases and deaths for percent black (third vs. first quartile) by urbanicity category

Demographic, underlying conditions, and social/ environmental variables, as well as COVID-19 cases and deaths are reported in Table 1 by varying proportions of black residents (<13% black residents versus ≥13% black residents). Counties with higher proportions of black residents experienced higher rates of COVID-19 cases and (in counties with >200 cases) deaths. Counties with higher proportions of black residents also had higher prevalence of comorbidities, proportions of individuals ≥65 years old, proportions of uninsured individuals, proportions unemployed persons; and higher air pollution.

In Table 2, the multivariate model examining predictors of both COVID-19 cases and associated mortality are presented. After controlling for potential confounders, a higher proportion of black residents in a county was associated with higher rates of COVID-19 cases (RR 1.24, 95% CI 1.17-1.33). Additionally higher rates of COVID-19 cases were independently associated with greater proportions of uninsured residents (RR 1.16, 95% CI 1.07-1.126), higher percentages of residents in crowded living conditions (RR 1.05, 95% CI 1.01-1.10), and more days since the first case (RR 3.1, 95% CI 2.9-3.3). Further, lower rates of COVID-19 diagnoses were associated with higher (i.e. poorer) social distancing scores (RR 0.88, 95% CI 0.84-0.92). The rate ratio associated with days since first case implies that counties with similar demographics and comorbidities tend to have about 3.1 times as many cases for each 15 additional days that COVID-19 has been detectable in the county.

**Table 2.** Adjusted^a^ Rate Ratios (Third Versus First Quartile) of Demographics’^b^, Chronic Disease^c^, Social/Environmental Factors^d^ with COVID-19 Diagnoses and Deaths

| Covariates | Inter-quartile range | Rate ratio COVID-19 cases (95% CI) | Rate ratio COVID-19 deaths (95% CI) |
| --- | --- | --- | --- |
| Percent black | (0.70, 10.30) | 1.23 (1.17, 1.33) | 1.18 (1.00, 1.40) |
| Percent white | (77.20, 95.10) | 1.02 (0.92, 1.13) | 0.93 (0.70, 1.23) |
| Percent over 65 years old | (14.48, 19.62) | 1.03 (0.97, 1.09) | 1.25 (1.08, 1.45) |
| Percent unemployed | (3.10, 4.80) | 0.89 (0.84, 0.94) | 0.95 (0.82, 1.11) |
| Percent uninsured | (7.51, 15.43) | 1.16 (1.07, 1.26) | 1.13 (0.91, 1.41) |
| Percent diabetes diagnoses | (9.30, 14.60) | 0.97 (0.92, 1.03) | 1.01 (0.88, 1.16) |
| Heart disease death rate | (154.40, 211.60) | 1.01 (0.96, 1.07) | 1.07 (0.91, 1.26) |
| HIV infection rate | (39.50, 192.00) | 1.00 (0.96, 1.04) | 1.01 (0.93, 1.10) |
| Cerebrovascular and hypertension death rate | (50.10, 89.00) | 1.02 (0.99, 1.05) | 1.03 (0.91, 1.11) |
| Urbanicity score | (3.00, 6.00) | 1.00 (0.92, 1.09) | 0.83 (0.66, 1.04) |
| Air toxins (PM<sub>2.5</sub>) | (4.58, 7.63) | 1.03 (0.93, 1.14) | 1.09 (0.87, 1.38) |
| Household occupancy >1 person per room | (1.23, 2.86) | 1.05 (1.01, 1.10) | 1.05 (0.93, 1.19) |
| Social distancing score | (3.00, 4.00) | 0.88 (0.84, 0.92) | 0.82 (0.73, 0.93) |
| Days since first case of diagnosis<sup>e</sup> | (12.00, 27.00) | 3.10 (2.89, 3.33) | 3.08 (2.50, 3.80) |

Note: rate ratios greater than one mean that higher levels of a given characteristic are associated with higher rates of COVID-19 cases or deaths
<sup>a</sup>Multivariable zero-inflated negative binomial regression model
<sup>b</sup>'Demographics' include percent black or white, percent over 65 years old, percent unemployed and percent uninsured, and household occupancy per room.
<sup>c</sup>'Chronic disease' includes diabetes diagnoses, heart disease deaths, cerebrovascular and hypertension deaths, and HIV diagnoses
<sup>d</sup>'Social/environmental' includes urbanicity (1=urban, 6=rural), PM<sub>2.5</sub> (fine particulate matter in the air), social distancing score is a grade ('A'=1, 'B'=2, 'C'=3, 'D'=4, 'F'=5)
<sup>e</sup>'Days since first case' is temporality variable based upon the following citation: Geographic Differences in COVID-19 Cases, Deaths, and Incidence — United States, February 12–April 7, 2020. MMWR Morb Mortal Wkly Rep 2020;69:465–471.
DOI: <http://dx.doi.org/10.15585/mmwr.mm6915e4external icon>
<sup>f</sup>Data as of April 13, 2020

For COVID-19 specific-mortality, after controlling for potential confounders, a higher proportion of black residents in a county was associated with higher rates of COVID-19 deaths (RR 1.18, 95% CI 1.00-1.40). Additionally, higher rates of COVID-19 deaths were associated with higher proportion of persons ≥ 65 years of age (RR 1.25, 95% CI 1.08-1.45), higher social distancing scores (RR 0.82, 95% CI 0.72-0.92), and longer length of time since the first case (RR 3.1, 95% CI 2.5-3.8).

In analyses of rates of COVID-19 cases and deaths stratified by level of urbanicity, the risks of COVID-19 diagnoses associated with higher black populations were similar across levels of urbanicity. The risks of COVID-19 death associated with higher black populations were not significant except in small metropolitan and non-core areas.

In the context of this analysis, population attributable fractions do not imply causality, but are used to depict excess cases associated with certain factors. Figure 4 shows that 280,112 cases of excess COVID-19 diagnoses were associated with occupancy of greater than one person per room, and 126,985 excess diagnoses of COVID-19 were associated with lack of health insurance. The population attributable fraction (PAF) for lack of health insurance was 3.3% for counties with <13% black residents and 4.2% for counties with ≥13% black residents. The protective effect of unemployment was also larger for counties with higher proportions of black residents. None of these determinants were associated with significant excess of COVID-19 deaths (data not shown).

**Figure 4.**
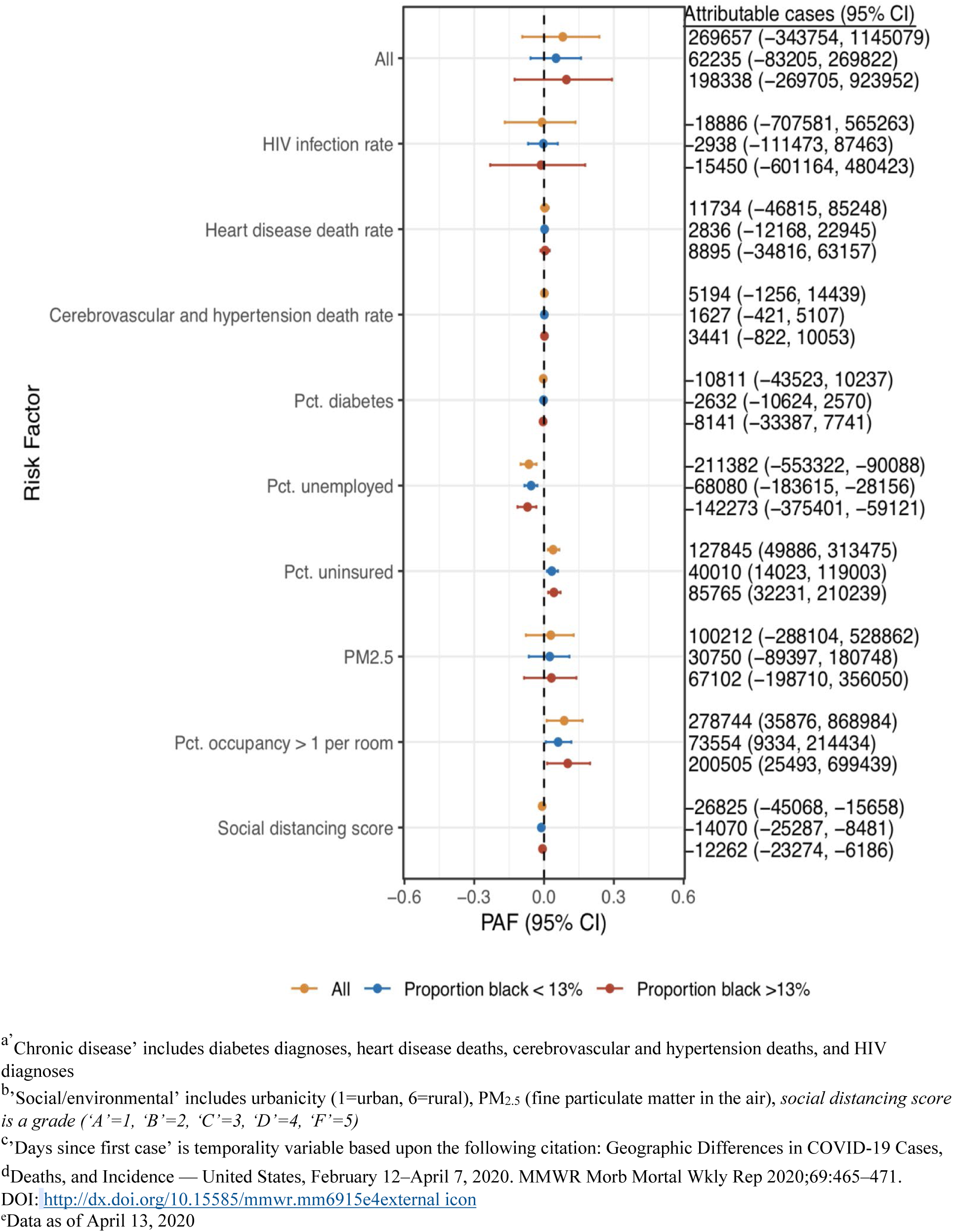
Estimated Number of COVID-19 Diagnoses Due to Chronic Disease^a^ and Social/Environmental^b^ Factors by Proportion of Black Residents

## Discussion

Collectively, these data demonstrate significantly higher rates of COVID-19 diagnoses and deaths in disproportionately black counties compared to other counties, as well as greater diabetes diagnoses, heart disease deaths, and cerebrovascular disease deaths in unadjusted analyses. Moreover, in the absence of complete national-level data disaggregated by race, county-level analyses offer an immediate alternative to measure the disproportionate impacts of COVID-19 diagnoses and deaths among black Americans. Importantly, our analyses indicated that disproportionate rates of COVID-19 cases and deaths persisted after controlling for potentially confounding factors that might be associated with both high rates of COVID-19 cases and deaths and with high proportions of black Americans. Roughly one in five US counties are disproportionately black and they accounted for five of ten COVID-19 diagnoses and nearly six of ten COVID-19 deaths nationally.

Greater health disparities in places with a greater concentration of black Americans is not unique to COVID-19. Similar patterns have been reported for other conditions such as HIV^27^, air pollution^28^, cancer^29^, and low birth weight^30^ and may be derived from the fact that in the United States, race often determines place of residence.^31^ Ninety-one percent of disproportionately black counties in these analyses are located in the southern United States – a region where most black Americans reside^32^ (58%) that also ranks highest in unemployment, uninsurance, and limited health system capacity or investment.^33^ These deficits are underscored by the finding that COVID-19 deaths in disproportionally black counties occurred at higher rates in rural and small metro counties.

Higher county-level unemployment was associated with fewer COVID-19 diagnoses. Employment presumably increases the likelihood of exposure to COVID-19, and this might differentially impact black Americans because only one in five black Americans has an occupation that permits working from home.^34^ Further, black Americans are overly represented in jobs that require both travel and regular interaction with the public, which can increase exposure to the virus, such as in the service industry (e.g. grocery store clerks, cashiers), transportation (e.g. bus drivers, subway train conductors), and health care (eg. nurses, medical aides, home healthcare workers).^35^ Being an ‘essential worker’ during the COVID-19 crisis carries risk, which is borne out in recent reports: CDC reported that over 9,000 health care workers nationwide have acquired COVID-19 and that black health care workers were disproportionately impacted (21% of infections; 13% of the population).^36^ Likewise, a report of New York City transit workers found more than 2,000 cases of COVID-19 and 50 deaths in a workforce that is 40% black, despite the black community comprising only a quarter of the New York City population.^37,38^

County-level lack of health insurance was associated with the proportion of black Americans in a county and with higher rates of COVID-19 diagnoses. Following the enactment of the Affordable Care Act (ACA), uninsurance rates fell by a third to 10.7% for black Americans from 2013 to 2016, but this rate climbed statistically significantly to 11.5% from 2016 to 2018.^39^ Nine of the 14 states that have not expanded Medicaid under the ACA are also located in the southern United States. Service industry jobs also have lower rates of insurance coverage than other professions.^35^ These associations point to the importance of policy-level interventions to eradicate interlocking inequities, but a review of interventions to address racial disparities found that policy interventions only accounted for 0.1% of all disparity interventions published over a thirty-year period.^40^

Disproportionately black counties were more urban than all other counties, were more likely to have >1 person per room which might reflect multigenerational and multifamily households, and marginally lower social distancing scores. Counties with more households having >1 person per room experienced greater rates COVID-19 cases, but counties reporting less social distancing had paradoxically fewer COVID-19 cases. These results may be due to the lack of granularity in the social distancing data when comparing counties rather than individuals, and potentially limited validation and imprecision of metrics used to gauge social distancing. Another explanation may be that metrics used to gauge social distancing could be biased because of differential access to personal mobile cellphones. A smaller percentage of households in nonurban metro and rural areas have mobile broadband access, 58.5% and 55.4%, respectively, compared to 68.2% for all households^41^; different patterns of phone sharing could result in estimates of social distancing biased towards less social interaction.

Leading public health experts have called for the rapid adoption of compulsory COVID-19 testing and surveillance reporting to include race/ethnicity, sex/gender, age, and educational level at the national, state, county, and zip code levels. Such reporting will be an important step to more rapidly and completely describing health inequities and informing programs.^42^ Deliberate attention to race and socio-economic barriers is needed when determining the locations of testing sites, yet an analysis in Philadelphia, where black people are a plurality of the population, found a 6:1 differential between testing in high-income versus low-income zip codes.^43^ Mitigating observed disparities among black communities could also be achieved by creating a special enrollment period for coverage under the ACA addressing suboptimal congregate living contexts including long term care facilities and homeless shelters, and remediating jail and prison overcrowding by appropriately releasing persons from confinement such as those charged but not convicted and others who may be at risk of severe illness if infected with COVID-19.^44^ Longer term, this crisis underscores the need for every state to expand Medicaid and for the federal and state governments to take coordinated actions to strengthen their insurance markets, expand health facility and provider capacity focusing on underserved populations and geographic areas, and greatly expand public investments in public health.

The results presented here should be interpreted in the context of several limitations. Given the aforementioned challenges in individual reporting of race in existing surveillance systems, the county-level data presented here represent an ecological analysis that could be subject to structural confounding where there are more black people in urban centers and urban centers have been more likely to be affected to date in the first wave of COVID-19. This possibility can be addressed by the responses to several questions. Are disproportionately black counties the urban counties affected early by COVID-19? The theory of structural confounding suggests that no individuals or low numbers of individuals in extreme cells (in this case, the most rural areas) raise concerns about the possibility of structural confounding.^45,46^ Our data reveal a trend to higher proportions of black population in the most urban counties. However, all five strata for lower urbanization still have 35%-50% of the percent black residents observed in the most urban counties. Within disproportionately black counties, is the impact of COVID-19 homogenous, or do relatively more black and less black areas within a given county have differential impact? Even within heavily impacted urban areas, predominantly black neighborhoods have higher rates of COVID-19 disease and death. For example, in Prince George’s County, Maryland, white suburbs are relatively unimpacted, while predominantly black suburbs are heavily impacted.^47^ Third, do excess COVID-19 diagnoses and deaths occur among black people in more rural counties, or in counties that experienced outbreaks later in the epidemic? The risk of COVID-19 diagnoses for counties with more black residents is consistent across levels of urbanicity. The risk of COVID-19-related death for disproportionately black counties is significantly higher only in small metro and rural areas. Further, where rural transmission has been significant, it has been in disproportionately black counties.^48,49^ Counties like Dougherty County Georgia experienced intense outbreaks among black Georgians a month later than the early wave of infections in large urban areas. In sum, the alignment of individual-level and neighborhood data within urban centers and the county-level data presented here documenting disproportionate burden among black people suggests that the ecologic analyses presented here are not attributable to structural confounding, and may hold true at the individual level in at least some cases.^49-53^

Although we have presented attributable fractions for associations of COVID-19 that are consistent with our conceptual framework^54^ of heterogeneity in the burden of disease and for secondary mortality, there is limited inference of causality in these relationships. Causal inference estimations are critical and represent a future direction for the analyses presented here. Importantly, COVID-19 transmission dynamics are complex and evolving and the inferences may evolve with additional data. We posit that these analyses of disproportionate burden among black people are likely conservative because black Americans are less likely to have health insurance and evidence is emerging across the country that black people with symptoms for COVID-19 are less likely than other groups to be tested for the virus. Ultimately, these analyses draw strength from highlighting specific counties at elevated risk for COVID-19 and identifying more locations with disparate COVID-19 outcomes among black Americans than those profiled in the media including Dougherty County, GA; St John the Baptist Parish, LA; Cook County, IL; Milwaukee County, WI; Queens County, NY.

Our paper focuses on black Americans, but they are not the only population of interest or at potentially elevated risk. Additional analyses exploring disparities in COVID-19 among Latino, Native American and other populations is critical as the results of inequitable outcomes are representative of past respiratory pathogens including H1N1^47^ in the U.S. and thus likely indicative of potentially future waves of COVID-19 and other rapidly emerging respiratory pathogens. Health disparities arise from a complex interplay of underlying social, environmental, economic, and structural inequities. We will continue to fail to address longstanding inequities until we commit to eliminating structural racism and the systemic roots that maintain and even reinforce these injustices.^55^ Ultimately, advancing the health and wellbeing of all Americans relies on leveraging these and other data to effect policy change that makes equity a reality in the U.S.

## Data Availability

All data used in the analysis are publicly available sources and are referenced to allow readers to access the public files.

